# The impact of housing insecurity on mental health, sleep and hypertension: a longitudinal analysis using linked UK household data, 2009-2019

**DOI:** 10.1101/2023.11.27.23299030

**Authors:** Kate E. Mason, Alexandros Alexiou, Ang Li, David Taylor-Robinson

## Abstract

**Background:** Housing insecurity is an escalating problem in the UK but there is limited evidence about its health impacts. Using nationally representative panel data and causally focussed methods, we examined the effect of insecure housing on mental health, sleep and blood pressure, during a period of government austerity.

**Methods:** We used longitudinal survey data (2009-2019) from the UK Household Longitudinal Study. Outcomes were probable common mental disorder, sleep disturbance due to worry, and new diagnoses of hypertension. The primary exposure was housing payment problems in the past year. Using doubly robust marginal structural models with inverse probability of treatment weights, we estimated absolute and relative health effects of housing payment problems, and population attributable fractions. In stratified analyses we assessed potentially heterogeneous impacts across the population, and potential modifying effects of government austerity measures. A negative control analysis was conducted to detect bias due to unmeasured confounding.

**Results:** Housing payment problems were associated with a 2.5 percentage point increased risk of experiencing a common mental disorder (95% CI 1.1%, 3.8%) and 2.0% increased risk of sleep disturbance (95% CI 0.7%, 3.3%). Estimates were larger for renters, younger people, less educated, households with children, and people living in areas most affected by austerity-related cuts to housing support services. We did not find consistent evidence for an association with hypertension (RD=0.6%; 95% CI -0.1%, 1.2%). The negative control analysis was not indicative of unmeasured confounding.

**Conclusions:** Housing payment problems were associated with worse mental health and sleep disturbance in a large UK sample. Households at risk of falling into rent or mortgage arrears need more support, especially in areas where housing support services have been diminished. Substantial investment is urgently needed to improve supply of social and affordable housing.

## INTRODUCTION

The threat or reality of being forced to move home is an important form of housing insecurity^1^, and an escalating problem in the UK. Rents relative to income are at their highest for a decade^2^, and rising interest rates are driving up mortgage costs, against a backdrop of stagnating wages, a freeze on housing benefit payments, and a cost-of-living crisis. Households under financial stress due to increased housing costs may forego non-housing essentials, or else fall behind with their rent or mortgage payments – and potentially both. Repeated failure to keep up with housing payments can lead to eviction for renters or home repossession for mortgage holders. England’s poorly regulated private rental sector has doubled since 2000, and the social housing sector is half the size it was in 1980^3^. Meanwhile, housing benefit payments for private renters have been cut and local authority spending on housing services declined by 40% as part of Government austerity measures following the financial crash of 2008-09^4^. This wider context leaves householders increasingly exposed to the socioeconomic, psychosocial, and physical risks of housing insecurity, including at the sharp end, homelessness.

Early in the COVID-19 pandemic, tenants were protected by an eviction moratorium, homeowners had access to mortgage support measures, the furlough scheme provided some income and employment protection for many, and welfare recipients had a temporary benefits uplift. With the withdrawal of these protections^5^, combined with rapid increases in the cost of living generally and housing in particular, a housing cost crisis has now emerged in the UK: the incidence of housing payment arrears is increasing, and eviction and repossession proceedings are on the rise^6,7^. Any population health burden attributable to these forms of housing insecurity should now also be expected to rise.

To better understand this health burden – and formulate appropriate policy and practice responses – requires more causally robust studies examining the health effects of the threat or actual experience of eviction or home repossession (or foreclosure in the United States) due to payment arrears. While a large body of evidence links various forms of housing disadvantage to poor mental and physical health outcomes,^8,9^ recent systematic reviews conclude that while several studies have examined the effects of foreclosure/repossession among mortgage holders^10^, the evidence relating to payment arrears and eviction of renters is limited, and most of the existing evidence base is centred on the United States^11,12^. Furthermore, drawing causal inferences from the existing evidence is hampered by likely time-varying confounding, the complex nature of the relationships involved, and a necessary reliance on observational data. A 2019 review^8^ of causally focussed studies of housing disadvantage (broadly defined) and mental health found only one focussed on the experience of falling behind in housing payments and one on eviction. In the UK, most studies of housing insecurity and health predate the large shifts in housing markets and the wider economy in the decade that followed the 2008-09 financial crash. An analysis of homeowners with a mortgage in the British Household Panel Survey (BHPS) reported evidence of a relationship between mortgage arrears and worsening mental health during the 1990-91 economic recession, with the association later weakening when the economic climate improved^13^. A 2009 study found that repossession of owned property was associated with increased risk of mental ill health, but no such relationship was apparent for eviction from a rental property^14^. A study of 27 European countries during the recent Great Recession reported a relationship between entering housing payment arrears and worse self-rated health in several countries including the UK^15^. Another study examined cross-sectionally (for the period 2010-12) the relationship between a range of housing characteristics and an inflammatory biomarker, with mixed findings^16^. All these studies carry a risk of bias due to time-varying confounding.

Our study addresses the dual need for an improved understanding of the relationship between housing insecurity and health during the post-recession decade of austerity in the UK, and for more robust studies of the health effects of the threat of losing one’s home. This evidence is needed to inform policy responses to the current housing and cost-of-living crises. We also examine unanswered questions about which aspects of health are most affected by the risk of losing one’s home, and what sociodemographic and contextual factors exacerbate or protect against negative health effects.

We consider three domains of health that are plausibly affected by housing insecurity: mental health, cardiovascular health, and sleep (Figure 1). Financial hardship is associated with poor mental health^17^ but we don’t know how much of the population’s mental health burden is attributable to housing insecurity, and there is limited high-quality evidence on this topic^8^. Cardiovascular health is linked to chronic stress^18^, but we know little about whether experiencing acute housing payment problems affects outcomes such as hypertension in the short term. And finally, sleep is emerging as an important influence on health^19^, with insufficient sleep linked to e.g. cardiovascular disease^20^, diabetes^21^ and depression^22^. Sleep disturbance due to worry is a likely proximal effect of housing insecurity, yet this has received limited research attention^23^.

**Figure 1.**
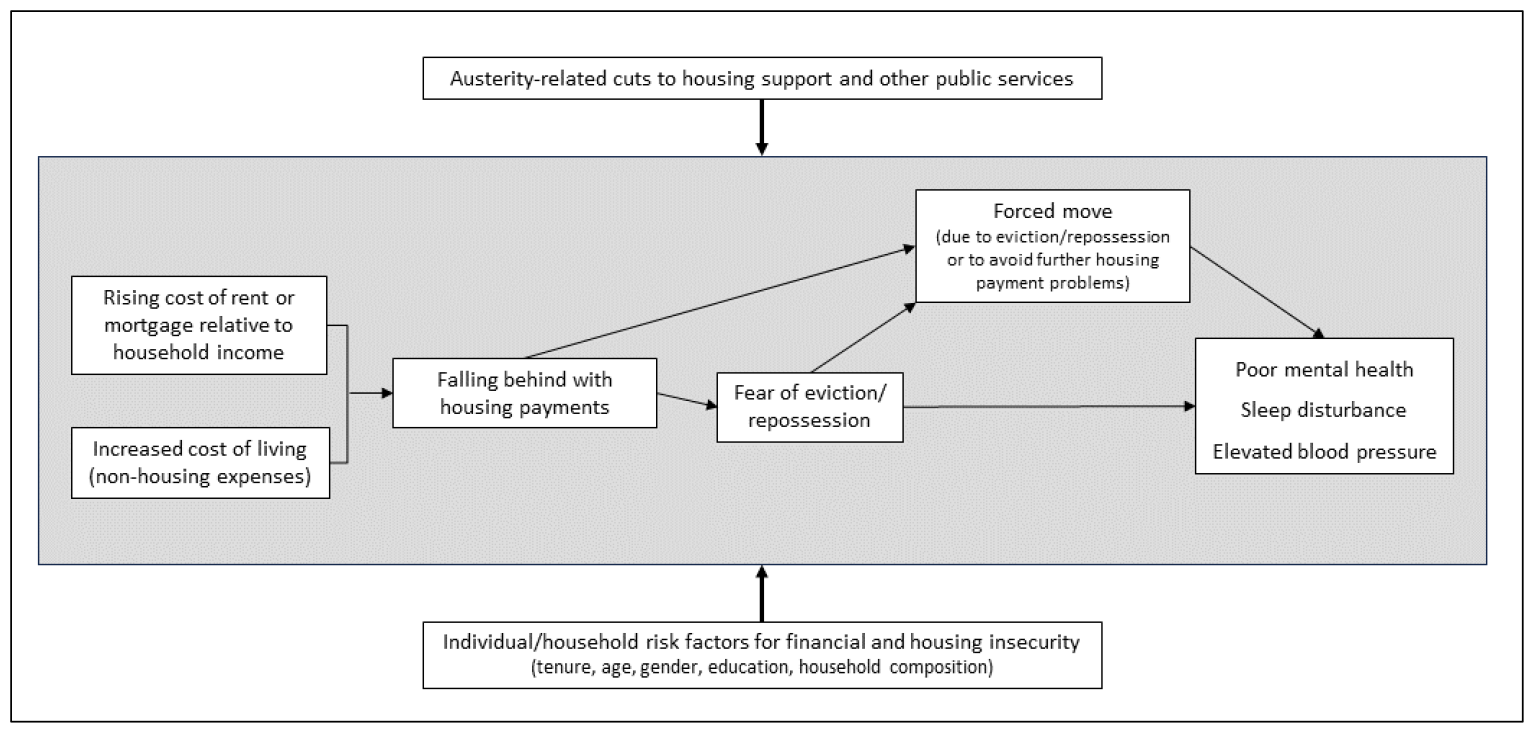
Logic model for pathways linking housing insecurity (housing payment problems and forced moves) to health, depicting potential modifying roles of austerity-related cuts to public services and individual and household factors. Note: This simplified model illustrates the primary relationships of interest in this study and the logic informing our analytical approach. See Figure S1 for a fuller causal diagram that includes confounding factors and pathways.

The aim of this study is to improve our understanding of the health consequences of housing insecurity in the UK during a period of government austerity and increasingly insecure housing market conditions. We applied marginal structural modelling with inverse-probability-of-treatment weighting, supplemented by a negative control analysis to detect possible residual confounding – methods better suited than most previous studies to estimating causal relationships. We asked: (1) does experiencing housing insecurity lead to poor mental health, sleep problems, and/or hypertension?; (2) do these relationships vary by other sociodemographic factors?; and (3) are these relationships stronger in places where austerity-related cuts to public services were deeper?

## METHODS

### Data and sample

We used survey data from nine annual waves of the UK Household Longitudinal Study (UKHLS) which began in 2009 and includes approximately 40,000 households randomly sampled from the UK population^24^. We used data spanning 2009-2019, which covers the introduction of austerity measures and the years following, while avoiding the COVID-19 pandemic period. We restricted analysis to working-age adults aged 25-64 years. We excluded young adults, as they are often living with parents and not primarily responsible for housing costs, and households without housing payment obligations (outright owners, households living rent-free).

To examine potential modifying effects of the UK government’s austerity policies of the 2010s, we linked the survey data to publicly available area-level data on local authority finance^25^ and public sector employment^26^. Due to limited data availability, this was only done for survey participants in England. Prior to linkage, we derived two area-level indicators of the severity of cuts to public services, based on annual measures of local authority (LA) expenditure on housing support services^27^, and public sector employment in each of the nine government office regions of England. The sharpest declines in both measures occurred between 2009 and 2012, so for each area we calculated the proportional change from its pre-austerity peak in 2009, to 2012. Thus, any observed effect modification relates to cuts occurring early in the study period, ensuring temporal precedence. For each area-level measure, we calculated the median change for England, classified each LA or region as above or below the median level, then linked these indicators to the UKHLS data via geographical identifiers for place of residence.

### Exposure

Our primary indicator of housing insecurity was self-reported housing payment problems, defined as living in a household that fell behind on rent or mortgage payments at any time in the past 12 months. As a secondary exposure we examined forced mobility, defined as having moved residence in the past 12 months either explicitly due to eviction or repossession, or any other move made by those who had reported difficulty paying rent or mortgage during the year^28^.

### Outcomes

#### Mental health

assessed using the 12-item General Health Questionnaire (GHQ-12), a validated screening tool for mental health problems in the general population. Participants rated the degree to which psychological symptoms were experienced in the last few weeks. A combined 0-12 score is then dichotomised such that ≥ 4 indicates probable common psychiatric morbidity^29^.

#### Sleep

Sleep disturbance is measured in each wave as part of the GHQ-12, with the question “Have you recently lost much sleep over worry?”. Responses on a 4-point scale from “not at all” to “much more than usual” were dichotomised to 0 = “not at all/no more than usual” and 1= “rather/much more than usual”.

#### Hypertension

At each survey wave, participants report any new diagnosis of high blood pressure since their previous interview.

#### Cancer (negative control)

At each survey wave, participants report any new diagnosis of cancer since their previous interview.

### Causal framework and potential confounders

Within a causal framework we sought to estimate the effect of housing insecurity on each health outcome. To inform our statistical analysis we used a directed acyclic graph (DAG) to depict our assumptions about the casual relationships between the variables of interest, based on existing literature and knowledge (Figure S1). This allowed us to identify the minimally sufficient set of covariates we need to account for to estimate the causal relationship between exposure and outcome. This set comprises the following confounders, some of which we conceptualise as time invariant and therefore include only baseline measures of, and others that vary over time and are included at each survey wave:

#### Time-invariant (baseline) confounders

self-reported gender, ethnicity, educational attainment [high: university degree; mid: upper secondary school; low: other/no formal qualification].

#### Time-varying confounders

age, age-squared, equivalised household income, employment status, household composition, long-term health condition, receipt of government benefits payments, geographical region, housing tenure.

Time-varying covariates were lagged by one year to ensure they preceded the exposure. We also included measures of income and employment in the current year and exposure and outcome status in the previous annual wave. Analyses for sleep and hypertension also included the lagged mental health measures as potential confounders. Models were adjusted for survey wave to account for time trends.

### Potential effect modifiers

We examined five housing or financial insecurity risk factors as potential effect modifiers: age group; tenure type; gender; household with/without dependent children; and educational qualifications. We also examined whether the local intensity of austerity policies acted as an effect modifier, by separately considering two distinct binary measures that capture different dimensions of austerity:

- per capita cuts to LA expenditure on housing support services (below or at/above England median)
- cuts to public sector employment (below or at/above England median)

We expect the first of these to be the stronger effect modifier, given its direct relevance to housing.

### Statistical methods

We used doubly robust marginal structural modelling (MSM) to estimate the effect of housing insecurity on each outcome. This involved using the confounders identified above to generate stabilised inverse-probability-of-treatment weights (IPTWs) for each individual at each survey wave, then applying these weights in adjusted logistic regression outcome models to achieve balanced distribution of potential confounders between exposed and unexposed individuals. The process of weighting individuals by their inverse probability of ‘treatment’ (in this case, exposure to housing insecurity) creates a pseudo-population in which exposure is independent of measured covariates, allowing us to obtain estimates of the average effect of interest. Under the assumption of no unmeasured confounding, these estimates can be interpreted as causal effects^30^. To assess whether weighting achieved sufficient balance of confounders, we calculated standardised mean differences between exposed and unexposed, and considered a difference ±0.1 SD to be statistically negligible. From each outcome model we estimated odds ratios, absolute risk differences and population attributable fractions, and their 95% confidence intervals, all in Stata 18.0 (see Supplementary Material for more details of statistical methods).

The analytical sample for the primary analyses consisted of 11,125 individuals (52,782 observations) for models of mental health, 11,164 individuals (52,948 observations) for sleep disturbance, and 8,125 individuals (38,196 observations) for blood pressure. Analysis of blood pressure diagnoses was restricted to respondents with no prior history of hypertension.

### Robustness checks

#### Negative control outcome analysis

Negative control outcome analyses are increasingly being used as a tool to detect residual confounding in observational studies, and thereby strengthen causal conclusions^31^. A negative control outcome is a variable that is not causally affected by the exposure of interest but is associated with the measured and unmeasured confounders that affect the relationship between the exposure and primary outcome(s)^32^. When, conditional on the same confounders measured in the primary analysis, an association is observed between the exposure of interest and a negative control outcome, this indicates the primary exposure-outcome association is biased due to inadequate control of confounding. The absence of any association between exposure and negative control outcome means there is no compelling empirical evidence of residual confounding in the primary analysis. In this paper, we employ any new diagnosis of cancer at time t, as a negative control outcome: it satisfies the two key assumptions of not being casually related to housing insecurity (at least not in the short time frame of our study) while at the same time being associated with the main measured confounders in our primary analysis (prior health, demographic and socioeconomic variables) and the potentially unmeasured confounders of that relationship (e.g. earlier life circumstances that may affect later health). In the negative control analyses, we substituted each primary outcome with self-reported cancer diagnosis by a health professional since the previous survey round, to check whether the expected null association was observed. As slightly different covariate sets and analytic samples were used for each of the three primary analyses, a separate negative control analysis was performed for each one.

#### Sensitivity analyses

We performed several additional analyses to check the sensitivity of our findings to various modelling choices:

1. To test sensitivity of our findings to the modelling approach, we ran conditional fixed-effects (FE) models as an alternative to the marginal structural modelling with IPTWs. While MSM is arguably more causally robust because it balances time-varying confounders across exposed and unexposed groups, FE models have the advantage of accounting for all time-invariant confounding, measured or unmeasured.
2. Inclusion of sampling weights in the calculation of IPTWs restricted the analytical sample to individuals with data for all waves – approximately half the available sample. For comparative purposes, we repeated the analysis without sampling weights.
3. The lead time to any hypothesised effect of housing insecurity on blood pressure is unknown, and potentially longer than for sleep and mental health. Additionally, a reported new diagnosis of high blood pressure in the past 12 months could have preceded the experience of housing insecurity. Therefore, for hypertension, we repeated the analysis lagging the exposure by one year (and the already lagged covariates by two years).

## RESULTS

The prevalence of having experienced housing payment problems in the past year was 11% in the analytical samples for mental health and sleep, and 10% in the sample for blood pressure. The prevalence of a forced move in any wave was 1%. The prevalence of a probable common mental disorder was 21%; 19% reported recently losing sleep due to worry; and 2% had received a new diagnosis of high blood pressure in the past 12 months (Table S1). A new diagnosis of cancer (the negative control outcome) was reported by 0.5% of the sample. Roughly half (54%) of all people who had ever experienced payment problems during the survey period did so in only one wave of follow up. Across the survey waves, between 31% and 43% of individuals who experienced housing payment problems in a given year had also reporting experiencing payment problems the previous year.

### Balance of confounding variables by inverse-probability-of-treatment weighting

Prior to weighting, most confounders of the primary relationships were substantially imbalanced between individuals exposed and unexposed to housing payment problems; gender, region, and age were the main exceptions. After IPT weighting, acceptable balance was achieved, with the standardised mean difference between exposed and unexposed within ±0.1 for all variables for the primary analyses (Figure 2). In the subgroup analyses, some standardised mean differences were slightly larger, but all were within ±0.2. For forced moves, however, IPTW did not achieve acceptable balance (Figure S2) so we did not progress to the MSM outcome model, and instead present only FE model results (Table S3).

**Figure 2:**
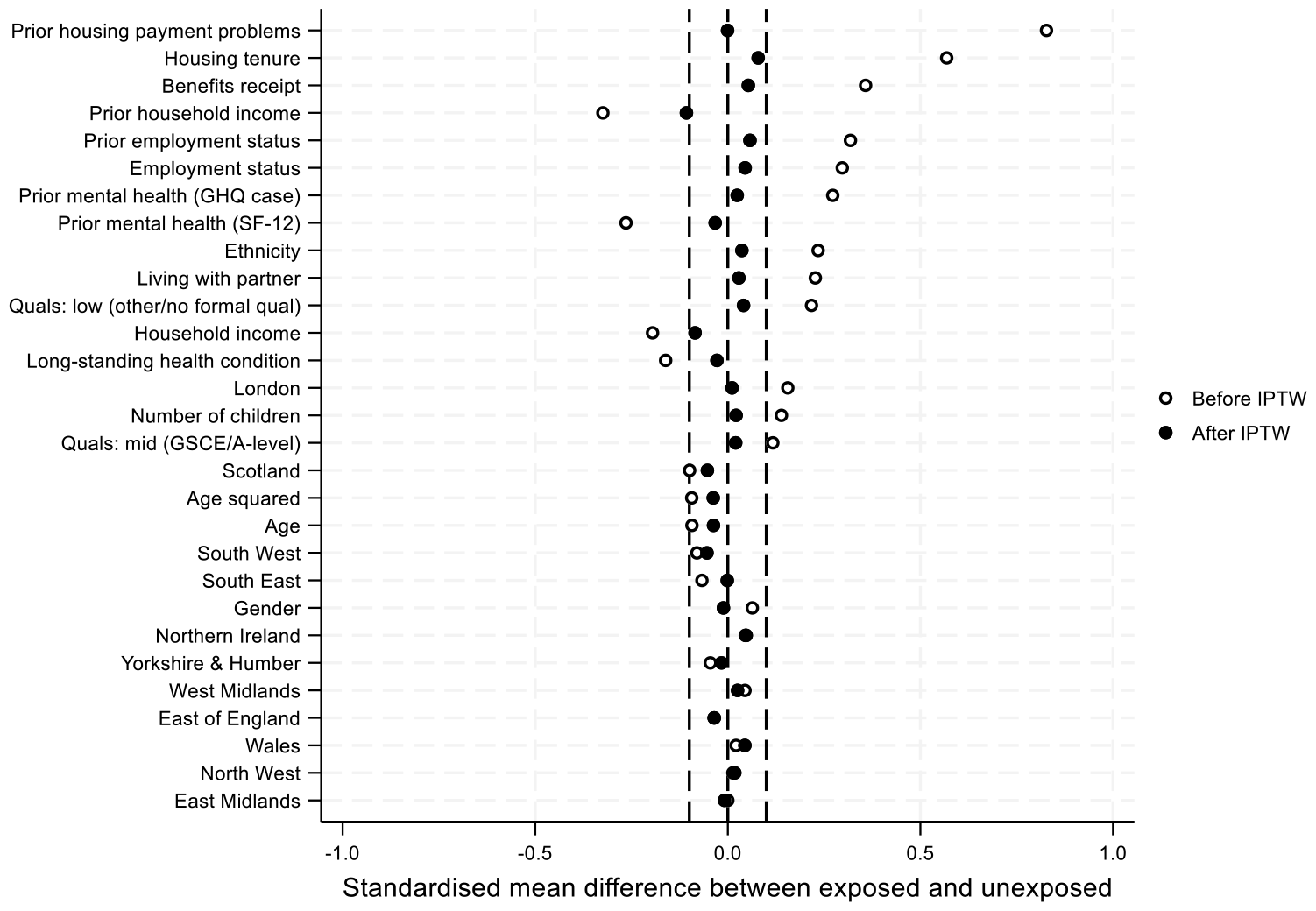
Housing payment problems: balance of confounder variables before and after inverse-probability-of-treatment weighting (IPTW) for primary analysis. Note: Standardised mean difference <0.1 (indicated by dotted vertical lines) was considered statistically negligible.

### Housing payment problems and health

The absolute difference in risk of developing a common mental health disorder comparing people who experienced housing payment problems in the past year with those who did not was 2.5 percentage points (95% CI 1.1%, 3.8%), with a corresponding odds ratio (OR) of 1.21 (95% CI: 1.09, 1.34) and a population attributable fraction (PAF) of 1.3% (95% CI: 0.6%, 2.0%), i.e. the results indicate 1.3% of the burden of poor mental health in the UK population aged 25-64 can be attributed to housing payment problems (Table 1). The risk of sleep disturbance due to worry was 2.0 percentage points higher for those who fell behind on their housing payments (95% CI: 0.7%, 3.3%), with a corresponding OR of 1.16 (95% CI: 1.06, 1.28) and a PAF of 1.1% (Table 1).

**Table 1.** Main results: association between exposure to housing payment problems and outcomes.

| Outcome | N individuals<br>(observations)<sup>a</sup> | Odds ratio<br>(95% CI) | Absolute risk<br>difference<sup>b</sup><br>(95% CI) | Population<br>attributable fraction<br>(95% CI) |
| --- | --- | --- | --- | --- |
| Probable common mental disorder | 11125 (52782) | 1.21 (1.09, 1.34) | 2.5% (1.1, 3.8) | 1.3% (0.6, 2.0) |
| Sleep disturbance due to worry | 11164 (52948) | 1.16 (1.06, 1.28) | 2.0% (0.7, 3.3) | 1.1% (0.4, 1.9) |
| New diagnosis of high blood pressure | 8125 (38196) | 1.36 (1.00, 1.84) | 0.6% (-0.1, 1.2) | 3.2% (-0.4, 6.7) |

<sup>a</sup> Sample sizes vary by outcome because of small differences in missing data across the outcomes, and for blood pressure the exclusion of individuals with a history of hypertension.
<sup>b</sup> Absolute risk difference is represented as percentage point difference between unexposed and exposed.

The estimated risk difference for new diagnoses of hypertension associated with housing payment problems was smaller, at 0.6 percentage points and the 95% confidence interval included no effect (−0.1%, 1.2%) (Table 1). Although the OR for hypertension was 1.36, the lower limit of the confidence interval was 1.00 and the PAF of 3.2% is very imprecise and may not necessarily represent a true effect (95% CI: -0.37, 6.69).

Stratifying by sociodemographic markers, we observed considerable effect heterogeneity. We found mental health and sleep to be more strongly related to housing payment problems the younger the age group, among private and social renters and social housing tenants, households with dependent children, and people without a university degree (Figures 3 and 4). Most notably we found housing insecurity to be associated with a risk difference for a common mental health disorder of 5.5% (95% CI: 2.3, 8.7) among private renters, and 4.6% (95% CI: 1.6, 7.6) for 25-34-year-olds (Figure 3a), and substantially elevated risk sleep disturbance due to worry for social housing tenants and people without a university degree after falling behind in housing payments (Figure 3b). In contrast, sleep disturbance and common mental health disorders showed little to no association with housing insecurity for people aged 55-64, those with a university degree, those without dependent children and homeowners with a mortgage. For new hypertension diagnoses, subgroup estimates lacked sufficient precision to draw conclusions (Figure S3).

**Figure 3.**
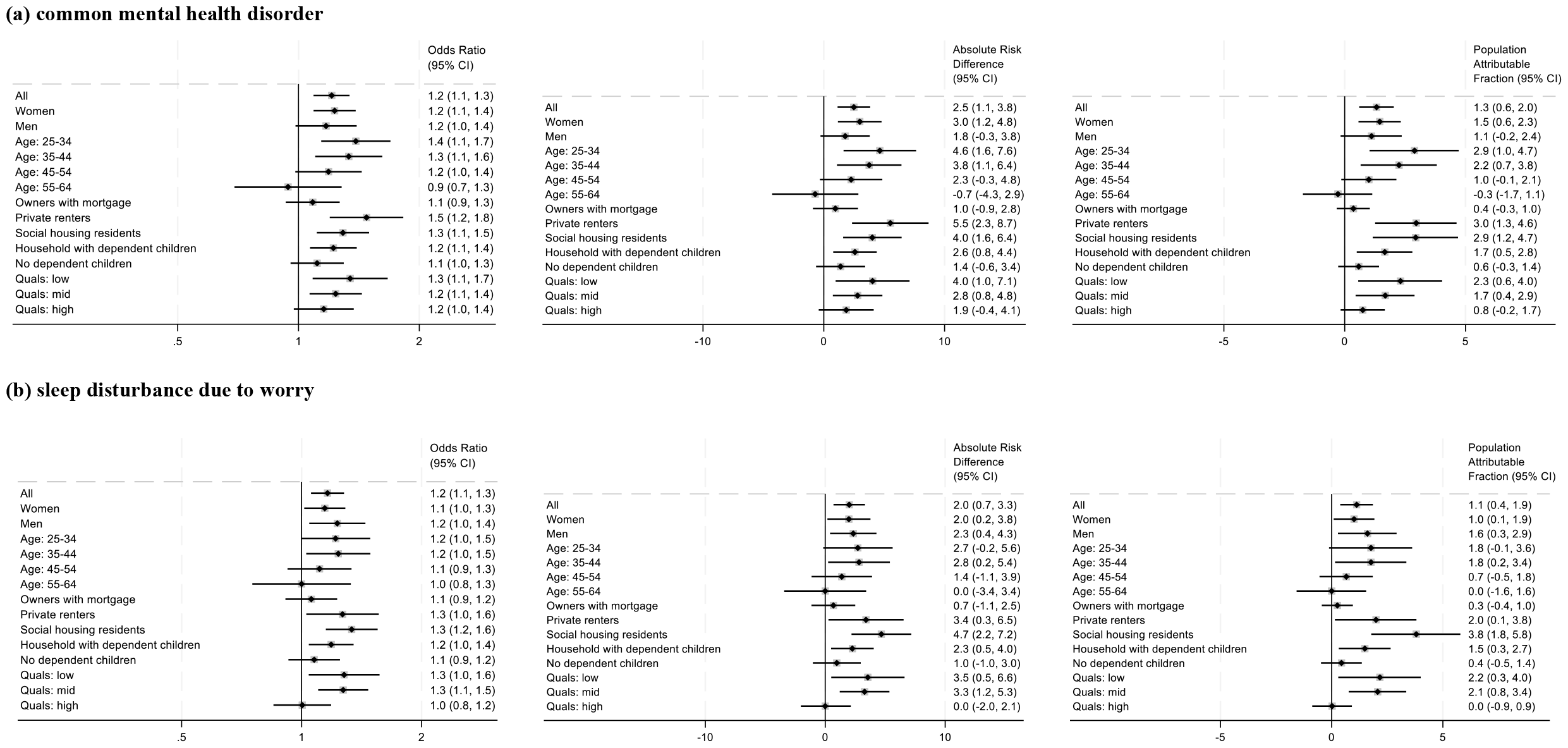
Estimated effects of housing payment problems on likelihood of (a) a common mental health disorder and (b) sleep disturbance due to worry, stratified by potential sociodemographic effect modifiers.

Overall effect estimates for England were slightly larger than for the whole of the UK (Tables 2, S2). People living in areas where cuts to LA spending on housing services had been greater, were more susceptible to poor mental health after experiencing insecure housing. The likelihood of experiencing a common mental disorder increased by 4.4% (2.2%, 6.6%) after falling behind in housing payments for people in areas with above-median cuts, and explained about 2.5% of the population prevalence of mental disorders in those areas, compared with statistically insignificant effects where spending cuts were less severe. There was no appreciable difference in the mental health effect of housing payment problems according to broader austerity indicator of public sector employment, nor was any significant effect heterogeneity by austerity measures observed for sleep disturbance (Table 2) or hypertension (Table S2).

**Table 2:** Estimated effects of housing payment problems on mental health and sleep disturbance, stratified by measures of intensity of austerity.

|  | N individuals<br>(observations) | Odds ratio<br>(95% CI) | Absolute risk<br>difference<br>(95% CI) | Population<br>attributable fraction<br>(95% CI) | P<sub>interaction</sub> |
| --- | --- | --- | --- | --- | --- |
| Common mental disorder |  |  |  |  |  |
| England (all) | 9412 (44866) | 1.22 (1.10, 1.36) | 2.7% (1.2, 4.1) | 1.4% (0.6, 2.21) |  |
| Cuts to LA housing services spending |  |  |  |  |  |
| above median level | 4451 (20937) | 1.39 (1.19, 1.62) | 4.4% (2.2, 6.6) | 2.5% (1.2, 3.7) | 0.016 |
| below median level | 4865 (23376) | 1.08 (0.93, 1.25) | 1.0% (-0.9, 3.0) | 0.5% (-0.5, 1.5) |  |
| Regional public sector employment cuts |  |  |  |  |  |
| >3% | 3628 (17362) | 1.24 (1.02, 1.49) | 2.8% (0.2, 5.3) | 1.3% (0.1, 2.5) | 0.593 |
| <3% | 5885 (27344) | 1.24 (1.08, 1.41) | 2.8% (1.0, 4.7) | 1.6% (0.6, 2.6) |  |
| Sleep disturbance |  |  |  |  |  |
| England (all) | 9348 (44455) | 1.19 (1.07, 1.32) | 2.31% (0.87, 3.76) | 1.31% (0.49, 2.12) |  |
| Cuts to LA housing services spending |  |  |  |  |  |
| above median level | 4468 (21007) | 1.26 (1.08, 1.48) | 3.14% (0.89, 5.38) | 1.85% (0.53, 3.15) | 0.648 |
| below median level | 4882 (23450) | 1.12 (0.98, 1.29) | 1.56% (-0.34, 3.47) | 0.84% (-0.19, 1.87) |  |
| Regional public sector employment cuts |  |  |  |  |  |
| >3% | 3641 (17427) | 1.14 (0.95, 1.36) | 1.70% (-0.69, 4.09) | 0.86% (-0.35, 2.05) | 0.823 |
| <3% | 5907 (27424) | 1.20 (1.06, 1.37) | 2.52% (0.67, 4.37) | 1.51% (0.41, 2.61) |  |

### Negative control analysis

No association was observed between housing payment problems and reporting a new diagnosis of cancer when this was used as a negative control for all three model instances (Table 3), providing support for causal inferences resting on the assumption that we have not omitted any important confounders of the exposure-outcome relationships of interest.

**Table 3.** Negative control analysis: odds ratios for the association between exposure to housing payment problems and new diagnosis of cancer in the same year (negative control outcome substituted for each of the three primary outcomes)

|  | Odds ratio (95% CI) |  |  |
| --- | --- | --- | --- |
|  | Replacing common<br>mental health disorder | Replacing sleep<br>problems due to worry | Replacing new diagnosis<br>of high blood pressure |
| Negative control: new diagnosis of cancer | 0.98 (0.58, 1.64)<br>p=0.937 | 0.99 (0.56, 1.66)<br>p=0.956 | 0.70 (0.35, 1.39)<br>p=0.306 |

### Sensitivity analyses

Results of sensitivity analyses are in Supplementary Table S4. To summarise, comparison of our primary models with alternative model specification indicates that estimates were broadly consistent across models and samples, and estimates from the primary models were among the most conservative.

When new blood pressure diagnoses were measured in the year after retrospective reporting of housing insecurity, rather than the same year, we saw no association with housing payment problems (OR=1.05, 95% CI: 0.75, 1.50, p=0.791).

## DISCUSSION

We found strong evidence that experiencing housing payment problems has a detrimental effect on mental health and sleep. Our estimates suggest that, all else being equal, the likelihood of developing a common mental health disorder is 2.4 percentage points higher for individuals who fell behind on their housing payment problems in the past year than those who were able to keep up with their payments. The risk of sleep disturbance due to worry was 2 percentage points higher. If these estimates represent true causal effects of insecure housing – which we discuss below – they translate to housing payment problems being responsible for 1.3% and 1.1% of the burden of poor mental health and sleep disturbance, respectively, in the UK working-age population. Evidence of an effect of housing payment problems on cardiovascular health was weaker.

For mental health and sleep disturbance, we consistently observed larger effect estimates for population subgroups who also tend to be more exposed to other forms of housing and financial insecurity: renters, younger people, those with fewer educational qualifications, and households with dependent children. The risk difference for a common mental health disorder increased to as much a 5.5% among private renters, and 4.6% for 25–34-year-olds, while social housing tenants and people without a university degree stand out as being particularly susceptible to sleep disturbance due to worry after falling behind in housing payments. In contrast, the mental health of older working-age people, mortgage holders, those with a degree, and those without dependent children was relatively protected when they faced housing payment problems. These results highlight the differential vulnerability to poor health outcomes of certain groups with less financial and housing security, and the role of housing costs in perpetuating health inequalities. We note that social housing residents and private renters appear to be similarly disproportionately susceptible to the mental health and sleep effects of housing payment problems compared to mortgage holders, despite social housing tenants having greater protections against eviction. This may be because housing payment problems are likely to follow a period of foregone spending on other essentials such as food or heating; thus, mental health and sleep impacts associated with falling behind on rent perhaps more accurately reflect the acute stress of not only the threat of losing one’s home, but additionally of other difficult trade-offs made trying to avoid rental arrears. The apparent protection of higher education, older age, home ownership and lack of dependent children may reflect these households’ greater flexibility in labour and housing markets and/or greater social capital on which to draw in times of financial stress.

These findings were observed during a period of austerity in the UK, characterised by cuts to housing support services, public sector employment, welfare and other public services. We hypothesised that any health effects of housing insecurity would be stronger in areas more severely impacted by austerity measures, and found some evidence to support this. We show that mental health effects of housing payment problems were indeed concentrated among people living in areas where expenditure on housing support services was cut most, whereas effects were felt across England regardless of the relative loss of public sector jobs in the wider region. This is consistent with a study of OECD countries that found high housing costs predicted more avoidable mortality and suicide in the years corresponding to our study period, than in the years before the financial crisis, and that some forms of government investment buffered against health harms of unaffordable housing^33^, and with findings from a British study of mortgage arrears and mental health before and after the 1990-91 recession^13^.

While we found only weak evidence at best of a short-term impact of housing insecurity on blood pressure, cardiovascular impacts of persistent housing insecurity may yet play out over the longer term. The immediate impact we observed of housing insecurity on sleep suggests this is plausible, given inadequate sleep has been linked to hypertension^34^.

### Strengths and limitations

A key strength of our study is the use of analytical methods better suited to causal inference than many earlier studies on this topic, contributing to an evidence base that otherwise lacks certainty. We applied a marginal structural modelling approach to a large, UK-representative longitudinal dataset to produce doubly robust effect estimates, supplemented by a negative control analysis to check for evidence of bias due to unmeasured confounding. Our negative control analysis showed the expected null association between housing insecurity and a new diagnosis of cancer in the same year, meaning there is no empirical indication that we have omitted (or poorly measured) important confounders of the exposure-outcome relationships of interest. This lends additional support for a causal interpretation of our primary findings. Our primary findings are also broadly robust to alternative model specifications, and are at the conservative end of the range of point estimates observed across our sensitivity analyses. This study also contributes new evidence from a period of recent UK history when housing markets and the wider economy underwent sizeable shifts linked not only to a global recession, but to a particular ideologically driven macroeconomic response to that recession, the public expenditure cuts of which were borne disproportionately by the least well off people and places in the UK, and which is now recognised as having been detrimental to social equity and health^35^.

Our causally focussed approach gives us greater confidence in a causal interpretation of our findings, however, we cannot entirely rule out the possibility of residual confounding, measurement error or selection bias. While our negative control analysis supports the assumption of no unmeasured confounding, especially with the point estimates from those analyses so close to the null value of one, our negative control outcome was rarer than our primary outcomes and the control analyses may therefore have been underpowered (particularly since cancer prevalence is much higher in older age groups, meaning cases will be rare in younger age groups). One plausible source of residual confounding is unmeasured, early-life circumstances not fully controlled for through more proximal, measured confounders. However, alongside our negative control analysis we are also reassured by the fact we observed larger, not smaller, effect estimates in the sensitivity analysis using fixed-effects models, which inherently control for baseline variation between people. FE models often yield conservative estimates due to their reliance on within-subject variation, yet for both mental health and sleep our primary MSM analysis produced the more conservative estimates. Another possible source of bias is measurement error in our coarse binary classification of some covariates, such as ethnicity, to avoid positivity problems arising from small numbers in some categories, and through the use of self-reported outcomes. There is also a risk of selection bias; for example, people more likely to be experience insecure housing and poor mental health or poor sleep might be underrepresented in the study data. We applied sampling weights to deal with attrition and baseline representativeness of the UKHLS sample, but some selection bias may remain. We observed differences between population subgroups, but we cannot entirely rule out the possibility that larger health effects among renters, younger adults and those with lower educational qualifications reflect more severe housing payment problems, on average, among those groups. While that might explain the observed effect heterogeneity, it nonetheless still highlights that these groups are most in need of targeted support, even if only because they are more exposed to housing insecurity rather than necessarily more susceptible to its health effects once exposed. Further research examining the intensity of housing insecurity is warranted to explore this distinction, and will become more important if housing cost pressures continue to escalate in the UK and elsewhere. The low prevalence of forced moves per survey wave is a further limitation of the study. Despite using a liberal definition of forced moves, numbers were small and this may have contributed to the failure to achieve confounder balance across exposed and unexposed individuals, precluding use of our preferred modelling strategy for that exposure.

### Implications and conclusions

The United Nations enshrines adequate housing as a basic human right, specifying that housing “is not adequate if its cost threatens or compromises the occupants’ enjoyment of other human rights”, including health^36^. Our study strongly suggests that housing insecurity has affected the health of many people in the UK, and that once exposed to housing payment problems, some population groups are especially susceptible to these health effects, as are people living in areas most affected by austerity-related cuts to housing support services. Our estimated effects are of similar magnitude to mental health effects of poverty recently published^37^. During our study period, 20% of individuals reported falling behind with their housing payments at least once. The current cost-of-living crisis means more people will be affected by housing payment problems in the coming months and years. A fifth of households in England and Wales currently live in private rental^38^, at a time when rent rises are outstripping wage increases, as landlords seek to recoup increased borrowing costs or, in some cases, simply take advantage of the market trends. Those forced to move are looking for a new home at a time of high demand^39^, adding to the psychological strain of finding suitable, affordable housing. Furthermore, the 17% of UK households in social housing don’t appear to be protected from the health impacts of housing payment problems, despite greater tenure security. Millions of UK mortgage-holders who took out fixed-rate home loans when interest rates were at record lows are due to shift to variable rates in the coming months, leaving them facing substantial increases in repayments. Whether this large group will be as protected from detrimental health effects as the owners in our pre-2019 sample were, remains to be seen.

With housing payment problems escalating, the health effects of housing insecurity pose a growing challenge on a large scale that requires recognition and policy action to protect population health and slow the widening of health gaps in UK society. In the short term, more support for households at risk of falling into arrears – particularly rent arrears – is needed, especially in areas where housing services have been diminished, as well as investment in mental health services. But tackling the root of the problem demands rapid and substantial investment to improve supply of social and affordable housing.

## Data Availability

UKHLS data are available to approved researchers through application to the UK Data Service. Local authority data are available online at https://pldr.org/ and https://webarchive.nationalarchives.gov.uk/ukgwa/20150506003720/http://www.ons.gov.uk/ons/rel/pse/public-sector-employment/regional-analysis-of-public-sector-employment--2012/index.html

## REFERENCES

1 Amore K, Baker M, Howden-Chapman P. The ETHOS definition and classification of homelessness: an analysis. Eur J Homelessness 2011; 5. https://www.feantsaresearch.org/download/article-1-33278065727831823087.pdf.

2 Rental Market Report: June 2023 - Zoopla. 2023; published online June 20. https://www.zoopla.co.uk/discover/property-news/rental-market-report/ (accessed June 21, 2023).

3 Ministry of Housing, Communities and Local Government. English Housing Survey: headline report. London, UK, 2020.

4 Alexiou A, Mason K, Fahy K, Taylor-Robinson D, Barr B. Assessing the impact of funding cuts to local housing services on drug and alcohol related mortality: a longitudinal study using area-level data in England. Int J Hous Policy 2021; 0: 1–19.

5 Winchester N. Universal credit: an end to the uplift. 2021; published online Sept 3. https://lordslibrary.parliament.uk/universal-credit-an-end-to-the-uplift/ (accessed June 27, 2023).

6 Mortgage and landlord possession statistics: July to September 2023. GOV.UK. 2023. https://www.gov.uk/government/statistics/mortgage-and-landlord-possession-statistics-july-to-september-2023/mortgage-and-landlord-possession-statistics-july-to-september-2023 (accessed Jan 23, 2024).

7 UK Finance. Mortgage Arrears Possessions Update - 18 May 2023. 2023. https://www.ukfinance.org.uk/data-and-research/data/arrears-and-possessions (accessed Aug 10, 2023).

8 Singh A, Daniel L, Baker E, Bentley R. Housing Disadvantage and Poor Mental Health: A Systematic Review. Am J Prev Med 2019; 57: 262–72.

9 Mansour A, Bentley R, Baker E, et al. Housing and health: an updated glossary. J Epidemiol Community Health 2022; published online June 27. DOI:10.1136/jech-2022-219085.

10 Tsai AC. Home Foreclosure, Health, and Mental Health: A Systematic Review of Individual, Aggregate, and Contextual Associations. PLOS ONE 2015; 10: e0123182.

11 Vásquez-Vera H, Palència L, Magna I, Mena C, Neira J, Borrell C. The threat of home eviction and its effects on health through the equity lens: A systematic review. Soc Sci Med 2017; 175: 199–208.

12 Chen KL, Miake-Lye IM, Begashaw MM, et al. Association of Promoting Housing Affordability and Stability With Improved Health Outcomes: A Systematic Review. JAMA Netw Open 2022; 5: e2239860.

13 Nettleton S, Burrows R. Mortgage Debt, Insecure Home Ownership and Health: An Exploratory Analysis. Sociol Health Illn 1998; 20: 731–53.

14 Pevalin DJ. Housing repossessions, evictions and common mental illness in the UK: results from a household panel study. J Epidemiol Community Health 2009; 63: 949–51.

15 Clair A, Loopstra R, Reeves A, McKee M, Dorling D, Stuckler D. The impact of housing payment problems on health status during economic recession: A comparative analysis of longitudinal EU SILC data of 27 European states, 2008–2010. SSM - Popul Health 2016; 2: 306–16.

16 Clair A, Hughes A. Housing and health: new evidence using biomarker data. J Epidemiol Community Health 2019; 73: 256–62.

17 Frankham C, Richardson T, Maguire N. Psychological factors associated with financial hardship and mental health: A systematic review. Clin Psychol Rev 2020; 77: 101832.

18 Steptoe A, Kivimäki M. Stress and cardiovascular disease. Nat Rev Cardiol 2012; 9: 360–70.

19 The Lancet. Waking up to the importance of sleep. The Lancet 2022; 400: 973.

20 Cappuccio FP, Cooper D, D’Elia L, Strazzullo P, Miller MA. Sleep duration predicts cardiovascular outcomes: a systematic review and meta-analysis of prospective studies. Eur Heart J 2011; 32: 1484–92.

21 Cappuccio FP, D’Elia L, Strazzullo P, Miller MA. Quantity and Quality of Sleep and Incidence of Type 2 Diabetes: A systematic review and meta-analysis. Diabetes Care 2009; 33: 414–20.

22 Zhai L, Zhang H, Zhang D. Sleep Duration and Depression Among Adults: A Meta-Analysis of Prospective Studies. Depress Anxiety 2015; 32: 664–70.

23 Bozick R, Troxel WM, Karoly LA. Housing insecurity and sleep among welfare recipients in California. Sleep 2021; 44: zsab005.

24 University of Essex, Institute for Social and Economic Research. Understanding Society: Waves 1-13, 2009-2022 and Harmonised BHPS: Waves 1-18, 1991-2009: Special Licence Access, Local Authority District. [data collection], 15th edn. UK Data Service, 2023 https://beta.ukdataservice.ac.uk/datacatalogue/doi/?id=6666#!#14 (accessed Nov 27, 2023).

25 Place-based Longitudinal Data Resource. https://pldr.org/ (accessed June 7, 2022).

26 Public Sector Employment, Regional Analysis of Public Sector Employment, 2012 ONS. https://webarchive.nationalarchives.gov.uk/ukgwa/20150506003720/http://www.ons.gov.uk/ons/rel/pse/public-sector-employment/regional-analysis-of-public-sector-employment--2012/index.html (accessed May 8, 2023).

27 Alexiou, Alexandros, Barr, Benjamin. Local Authority Finance: Gross Current Expenditure - Housing services (GFRA only) (FIN_07_25). 2021. doi:10.17638/datacat.liverpool.ac.uk/1368.

28 Ong ViforJ R, Hewton J, Bawa S, Singh R. Forced housing mobility and mental wellbeing: evidence from Australia. Int J Hous Policy 2023; 23: 138–62.

29 Jackson C. The General Health Questionnaire. Occup Med 2007; 57: 79.

30 Hernan MA, Robins JM. Causal Inference: What If. Boca Raton: Chapman & Hall/CRC, 2020 https://www.hsph.harvard.edu/miguel-hernan/causal-inference-book/ (accessed Aug 23, 2022).

31 Shi X, Miao W, Tchetgen ET. A Selective Review of Negative Control Methods in Epidemiology. Curr Epidemiol Rep 2020; 7: 190–202.

32 Lipsitch M, Tchetgen Tchetgen E, Cohen T. Negative controls: a tool for detecting confounding and bias in observational studies. Epidemiology 2010; 21: 383–8.

33 Park G-R, Grignon M, Young M, Dunn JR. The association between housing cost burden and avoidable mortality in wealthy countries: cross-national analysis of social and housing policies, 2000-2017. J Epidemiol Community Health 2023; 77: 65–73.

34 Meng L, Zheng Y, Hui R. The relationship of sleep duration and insomnia to risk of hypertension incidence: a meta-analysis of prospective cohort studies. Hypertens Res 2013; 36: 985–95.

35 Schrecker T, Bambra C. How politics makes us sick: neoliberal epidemics. London: Palgrave Macmillan, 2015.

36 UN Habitat. The Right to Adequate Housing: Fact Sheet No. 21 (Rev. 1). 1993. https://www.ohchr.org/en/publications/fact-sheets/fact-sheet-no-21-rev-1-human-right-adequate-housing (accessed June 29, 2023).

37 Thomson RM, Kopasker D, Leyland A, Pearce A, Katikireddi SV. Effects of poverty on mental health in the UK working-age population: causal analyses of the UK Household Longitudinal Study. Int J Epidemiol 2022;: dyac226.

38 Housing, England and Wales: Census 2021. https://www.ons.gov.uk/peoplepopulationandcommunity/housing/bulletins/housingenglandandwales/census2021#tenure (accessed June 29, 2023).

39 Renters compete with 20 others in battle to find a home. BBC News. 2023; published online July 25. https://www.bbc.com/news/business-66246223 (accessed Aug 8, 2023).

