## Supplementary material for "The impact of housing insecurity on mental health, sleep and hypertension: a longitudinal analysis using linked UK household data, 2009-2019"

### Area-level data on local authority finance and regional public sector employment

To examine potential modifying effects of cuts to public services related to the UK government's austerity policies of the 2010s, we linked UKHLS survey data to area-level data on local authority finance and regional public sector employment. The local authority finance data derive from publicly available, annual revenue outturn estimates of Local Authority (LA) revenue expenditure and financing, and have been compiled into panel data available in our Place-based Longitudinal Data Resource[25]. We used the measure of total funding distributed from central government to each LA each year[26], and the measure of annual LA expenditure on housing support services[27] (which reflects local prioritisation influenced by housing-related and competing need, and constrained by the funding available both from central government grants and council tax). Both were adjusted for inflation[28] and converted to per capita values. The sharpest decline in funding and expenditure occurred in the period up to 2012, so for each LA we calculated the 2012 value of each finance measure as a proportion of the value in the year of its pre-austerity peak (2009 for housing support services and 2010 for central funding), to represent the severity of austerity related cuts affecting the LA. Defining our austerity measures at 2012 means that any observed effect modification is related to the cuts occurring early in the study period, thus ensuring temporal precedence. At the level of the nine government office regions of England, we used data from the Office for National Statistics on public sector employment[29] to calculate the change in the number of public sector employees in each region between Q3 2009 and Q3 2012. From each of these derived area-level datasets, we calculated the median value for England and identified whether each LA or region was above or below the median level, and then linked these indicators to the survey data, via identifiers for the administrative region and local authority in which each UKHLS participant resided at each survey wave.

### Statistical methods

*Inverse probability of treatment weighting (IPTW) and balance checking*

IPTWs were generated by predicting the probability of each individual experiencing each housing insecurity variable, conditional on all time-invariant and time-varying confounders, dummy variables for each survey wave, and survey weights to deal with sample attrition and ensure the sample better reflected the UK-representative sample recruited at baseline. These predicted probabilities were then stabilized by using predicted values from an empty logistic model of each housing insecurity measure as the numerator of the IPTW. To remove the influence of extreme weights, the stabilised IPTWs were truncated at the top and bottom 1000$^{th}$ centiles.

We calculated standardised mean differences between exposed and unexposed to assess whether sufficient balance had been achieved, with a difference between -0.1 and 0.1 considered statistically negligible.

*Outcome models*

Once derived, the IPTWs were included as sampling weights in pooled logistic regression models estimating the odds ratio for each exposure-outcome combination. We included the full set of potential confounders as covariates in these outcome models as well as the IPTW models, an approach referred to as 'doubly robust' because only one or other of the outcome model or the model to generate the IPTWs need be correctly specified for the estimator to be unbiased[32]. Standard errors were adjusted for clustering within individuals over time. Using post-estimation commands in STATA we also estimated the absolute risk difference and population attributable fractions from each model, alongside the odds ratios and 95% confidence intervals.

### Secondary analysis: forced moves and health

*Definition of exposure*

As a secondary exposure we examined forced mobility, defined as having moved residence in the past 12 months either explicitly due to eviction or repossession, or any other move made by those who had reported difficulty paying rent or mortgage during the year. This definition of forced move captures residential moves made to avoid ongoing payment problems or formal eviction due to arrears[30]. Although it risks misclassifying some moves coincident with but unrelated to payment problems, we chose this broader definition due to the small number of evictions in the sample.

*Balance of confounding variables by inverse-probability-of-treatment weighting*

For forced moves, IPTW did not achieve acceptable balance of confounders between individuals exposed and unexposed to a forced move (Supplementary Figure 2). Therefore we did not progress to the MSM outcome model. Instead we estimated conditional fixed effects models, equivalent to those used in Sensitivity Analysis 1 for housing payment problems, recognising that these estimates are likely to be biased by unmeasured time-varying confounders (but not by time-invariant confounding, due to the nature fixed effects models which estimate within-person change and therefore account fully for any baseline differences between individuals).

*Results*

Compared to those who did not experience a forced move, those who did showed higher odds of a common mental health disorder (OR 1.67, 95% CI: 1.14, 2.45).  and sleep disturbance (OR 1.50, 95% CI: 1.02, 2.20) (Supplementary Table 1). Contrasting these results with the equivalent FE models for housing payment problems (Supplementary Table 2) suggests, as would be expected, that actually being forced to move may have a larger effect on mental health and sleep than only the risk of a forced move. However, the persistence of substantial confounder imbalance limits our ability to interpret these estimates using a counterfactual framework. For hypertension, the sample size was greatly reduced and the OR was imprecise and not statistically significant (OR=0.55, 95% CI: 0.18, 1.65).

**Robustness checks/sensitivity analyses**

In addition to the negative control analysis reported fully in the main text, we also checked the robustness of our findings by undertaking the following sensitivity analyses:

1. To test the sensitivity of our findings to the modelling approach, we also ran conditional fixed-effects (FE) logistic regression models as an alternative to the marginal structural modelling with IPTWs approach. While MSM with IPTW is arguably more causally robust because it explicitly balances time-varying confounders across exposed and unexposed groups, the advantage of FE models over the MSM approach is that they account for all unmeasured time-invariant confounding, for example earlier life circumstances that may affect risk of housing insecurity and health outcomes.

2. Inclusion of UKHLS survey weights in the calculation of IPTWs restricts the analytical sample to individuals with survey data for all waves, which is only about half the available sample. For comparative purposes, we repeated the analysis without using the sampling weights in the IPTW estimation, on the same restricted sample, and then on the larger, unrestricted sample.

3. The lead time to any hypothesised effect of housing insecurity on blood pressure is unknown, and may be longer than for sleep and mental health, which could be expected to be immediate. Additionally, while the nature of the sleep and mental health variables were such that their temporal sequencing was more or less assured as being after the exposure, a reported new diagnosis of high blood pressure in the past 12 months means it could have preceded the experience of housing insecurity. Therefore, for hypertension, we repeated the analysis after lagging the exposure by one year (and the already lagged covariates by two years), i.e. we use the blood pressure diagnoses measures from t+1.

*Results of sensitivity analyses*

Results of sensitivity analyses are shown in Supplementary Table 4. For all three outcomes, comparison of our primary models with alternative model specification indicates that estimates were broadly consistent across models and samples, and estimates from our primary models were among the most conservative. Point estimates from FE models were uniformly larger and less precise than MSM models. Compared with the primary analyses where survey weights were incorporated into the calculation of the IPTWs to account for sample attrition, the MSM model on the same sample but not incorporating survey weights yielded almost identical results. For mental health and sleep, relaxing the restriction of the sample imposed by the inclusion of survey weights, to no longer require individuals to have data at all waves, generated more precise estimates, but also appeared to drive point estimates slightly away from the null.

**Figure S1: Directed acyclic graph (DAG) of the relationship between housing insecurity and health outcomes.** Note: for simplicity this shows only two timepoints

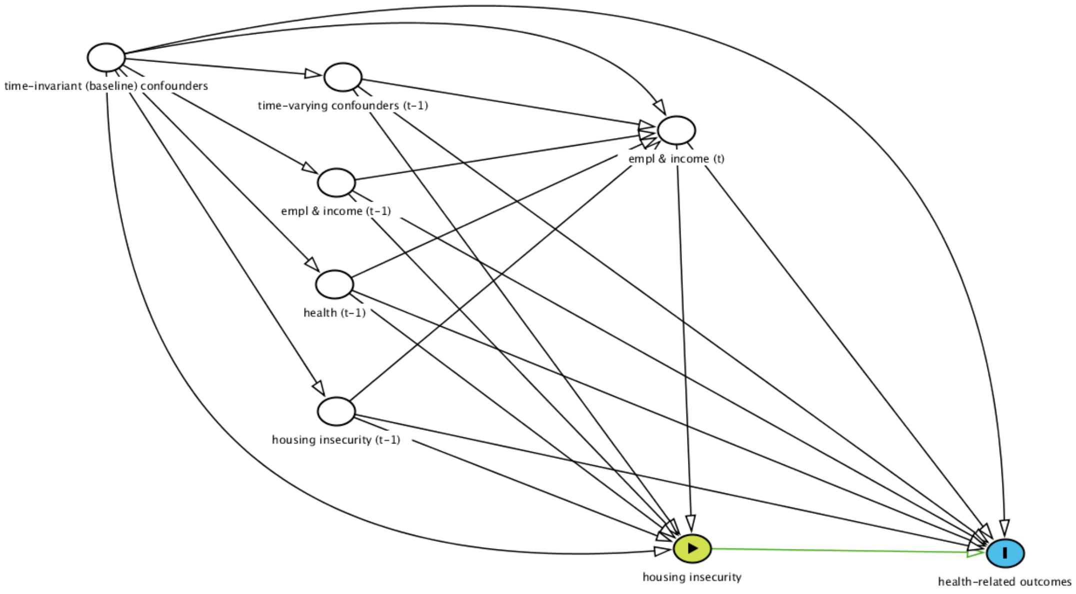


**Table S1. Sample sizes and exposure and outcome prevalence for pooled analytical samples**

| | Mental health analysis | Sleep analysis | Blood pressure analysis |
|---|---|---|---|
| Sample size (individuals (observations)) | 11,125 (52,782) | 11,164 (52,948) | 8,125 (38,196) |
| Housing payment problems in past 12 months (%) | 10.6 | 10.6 | 10.2 |
| Probable common mental health disorder (%) | 20.5 | na | na |
| Sleep disturbance due to worry in recent weeks (%) | na | 19.3 | na |
| New diagnosis of high blood pressure in past year (%) | na | na | 1.8 |
| New diagnosis of cancer in past year (%; negative control) | 0.5 | 0.5 | 0.5 |

**Figure S2: Forced moves analysis: balance of confounder variables before and after inverse probability of treatment weighting for primary analysis.**

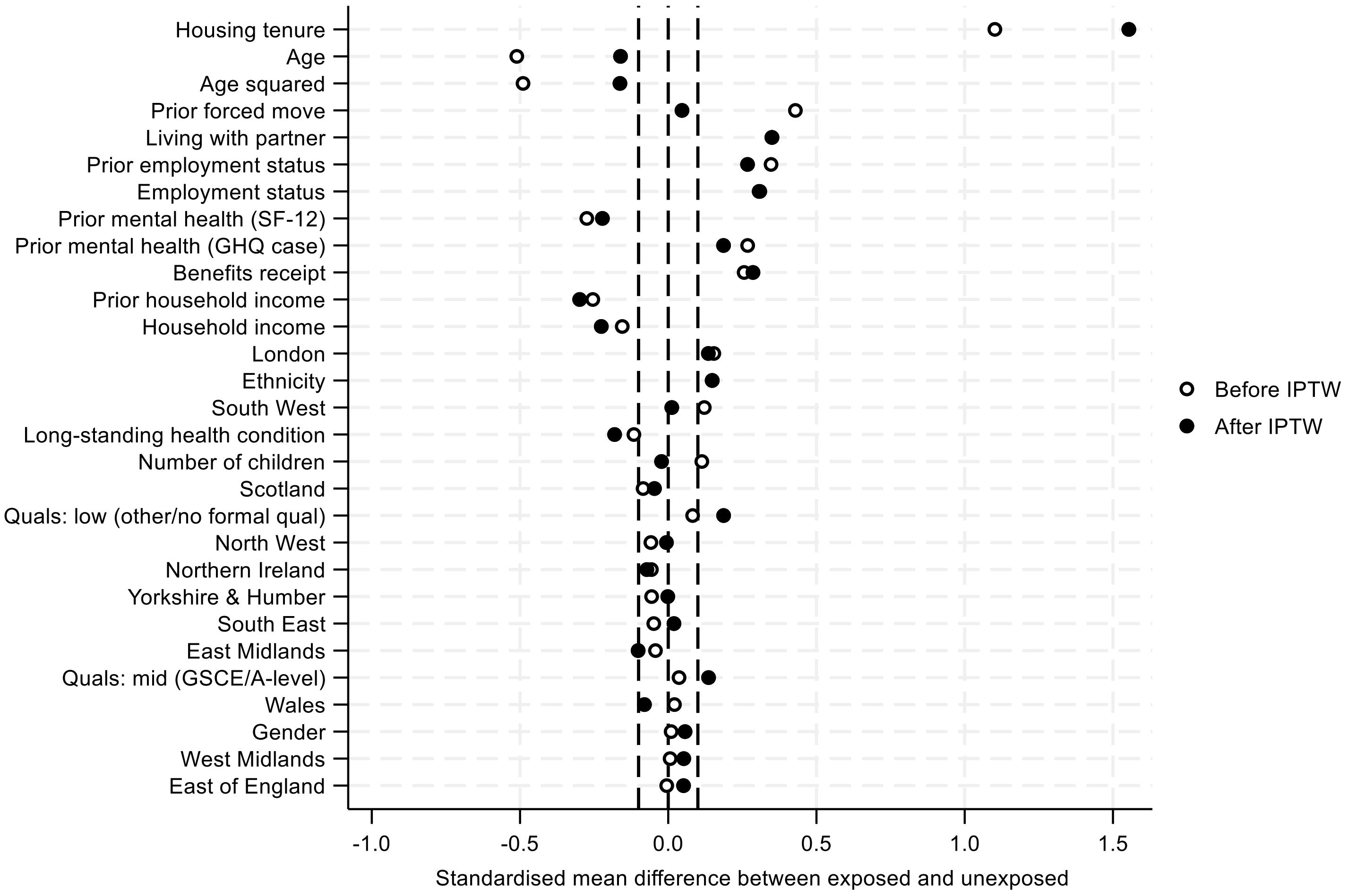

**Figure S3. Estimated causal effects of housing payment problems on likelihood of a new diagnosis of high blood pressure, stratified by sociodemographic potential effect modifiers**

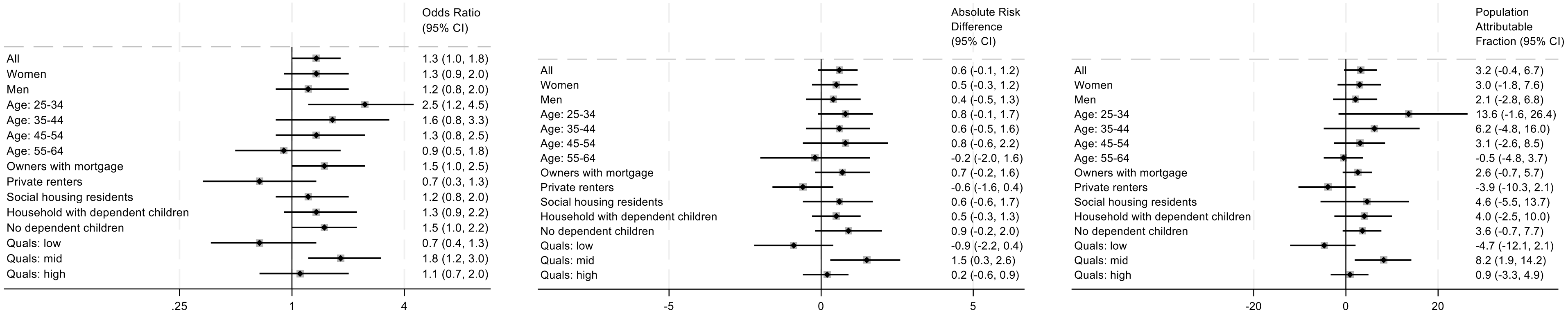


**Table S2: New blood pressure diagnosis estimates stratified by measures of austerity intensity**

| Subgroup | N individuals (observations) | Odds ratio (95% CI) | Absolute risk difference (95% CI) | Population attributable fraction (95% CI) | $P_{interaction}$ |
|---|---|---|---|---|---|
| England (all) | 6835 (32142) | 1.51 (1.08, 2.09) | 0.74% (0.06, 1.43) | 4.54% (0.35, 8.56) | |
| *Cuts to LA housing services spending* | | | | | |
| above median level | 3216 (15088) | 1.56 (1.01, 2.41) | 0.88% (-0.13, 1.89) | 5.18% (-0.76, 10.77) | 0.923 |
| below median level | 3620 (17054) | 1.50 (0.94, 2.42) | 0.67% (-0.23, 1.57) | 4.26% (-1.54, 9.72) | |
| *Regional public sector employment cuts* | | | | | |
| >3% | 2729 (12783) | 0.86 (0.47, 1.56) | -0.21% (-1.01, 0.59) | -1.17% (-5.66, 3.13) | 0.135 |
| <3% | 4272 (19666) | 1.71 (1.17, 2.48) | 1.03% (0.17, 1.88) | 6.76% (1.15, 12.06) | |

**Table S3: Results from conditional fixed effects logistic regression models for forced moves and mental health, sleep and blood pressure**

| | *Probable common mental disorder* | | *Sleep disturbance due to worry* | | *New diagnosis of high blood pressure* | |
|---|---|---|---|---|---|---|
| | Number of individuals (N observations) | Odds ratio (95% CI) | Number of individuals (N observations) | Odds ratio (95% CI) | Number of individuals (N observations) | Odds ratio (95% CI) |
| Forced move (FE model) | 3468 (19915) | 1.67 (1.13, 2.45) | 3590 (20571) | 1.50 (1.02, 2.20) | 475 (2711) | 0.55 (0.18, 1.65) |

Note: Models adjusted for equivalised household income, employment status, household composition, long-term health condition, receipt of government benefits payments, geographical region, housing tenure, all lagged by one year to ensure they preceded the exposure. Models also included age, age-squared, survey wave, prior outcome status, and income and employment in the current year. Analyses for sleep and hypertension also included lagged mental health.

**Table S4: Results of sensitivity analyses for comparison with primary MSM results (presented in Table 1)**

| | *Probable common mental disorder* | | *Sleep disturbance due to worry* | | *New diagnosis of high blood pressure* | |
|---|---|---|---|---|---|---|
| | Number of individuals (N observations) | Odds ratio (95% CI) | Number of individuals (N observations) | Odds ratio (95% CI) | Number of individuals (N observations) | Odds ratio (95% CI) |
| FE with survey weights | 4004 (25169) | 1.34 (1.18, 1.53) | 4177 (26169) | 1.37 (1.21, 1.55) | 467 (2667) | 1.49 (1.02, 2.18) |
| MSM without survey weights (unrestricted* sample) | 29076 (114974) | 1.24 (1.16, 1.32) | 29173 (115348) | 1.23 (1.16, 1.31) | 24204 (93733) | 1.25 (1.02, 1.51) |
| MSM without survey weights (restricted* sample) | 11126 (52782) | 1.22 (1.10, 1.34) | 11164 (52948) | 1.15 (1.05, 1.27) | 8125 (38196) | 1.40 (1.04, 1.90) |
| Hypertension outcome at t+1 | n/a | n/a | n/a | n/a | 7128 (30076) | 1.05 (0.73, 1.50) |

Notes: Sample sizes for FE models are much smaller than MSM because individuals are excluded when there is no variation in the outcome over their full set of observations. FE models are adjusted for only time-varying confounders because these are within-person models and so automatically control for any time-invariant confounders.

*Restricted to only those individuals with data for every survey wave, i.e. same sample as the primary analysis
